# Product Features Used to Promote Top-selling Cannabis Vape Products in An Online Retail Environment

**DOI:** 10.1101/2024.11.13.24317258

**Authors:** Julia Chen-Sankey, Kathryn La Capria, Siyan Meng, Rosanna Mazzeo, Neha Vijayakumar, Alisa A. Padon, Kimberly G. Wagoner, Meghan B. Moran, Matthew Rossheim, Carla Berg, Kristina M. Jackson

**Author notes:** **Corresponding author:** Julia Chen-Sankey, PhD, MPP, Rutgers School of Public Health, 303 George St. Room 525, New Brunswick, NJ 08901.

## Abstract

**Introduction:** Exposure to commercial marketing of cannabis vape products (CVPs) may encourage CVP use, yet little is known about how these products are marketed. This study examined marketing descriptions of CVPs’ functions and benefits in an online retail environment.

**Methods:** Product descriptions from a sample of 343 CVPs from 78 top-selling cannabis brands in 2023 were obtained from a large cannabis e-commerce website. Each description was thematically coded based on the promoted product features.

**Results:** The most frequently mentioned product feature was flavor profile and sensation (74.1%), including general and specific flavor descriptors (e.g., fruit flavors) and sensory experiences (e.g., sweet, spicy). Psychoactive effects were noted in 47.5% of the descriptions, detailing potency or effects such as feeling “high,” “stoned,” or “buzzed.” Product quality, such as “purity,” “natural,” or “organic,” was mentioned in 42.3% of the descriptions. Other product benefits described included mood enhancement (33.8%), reduced harm (24.2%), relaxation/tension reduction (23.9%), therapeutic effects (18.4%), focus/creativity (16.6%), convenience/discreetness (11.1%), socialization enhancement (6.7%), and physical performance enhancement (6.1%).

**Discussion:** Some frequently promoted product features of CVPs, such as flavor and sensation and psychoactive effects, may particularly appeal to younger consumers for non-medical use. Meanwhile, attributes surrounding high product quality and reduced harm may reduce individuals’ perceived risks of CVP use and may violate state restrictions on therapeutic claims. Additional research assessing the influence of exposure to marketed product features on product perceptions and use behaviors among priority populations is needed to inform regulations and enforcement.

## INTRODUCTION

As of 2024, more than 20 states in the U.S. have legalized non-medical cannabis.^1^ Moreover, a 2023 study showed that regular delta-9 THC products for smoking or vaping were being sold in 61% of smoke shops in states without legal non-medical cannabis retail.^2^ There are many different types of cannabis products, which are consumed in different ways, including combustibles (for smoking), vapes, edibles, topicals, tinctures, and ingestible pills/capsules, etc.^3^ The past few years have seen tremendous growth in cannabis vape products (CVP) used to inhale and exhale an aerosol or vapor made from cannabis concentrates or dry flowers that are heated in an electronic-powered device.^4^ In 2023, the sales of CVPs constituted 33% of the cannabis market in the U.S., ranking second after flower,^5^ and such market share is projected to increase over the next few years.^6,7^

CVP use is especially popular among youth and young adults in the U.S.^8–10^ In 2023, 14% and 20% of 12^th^ graders reported past 30-day and past-year cannabis vaping, respectively.^11^ In the same year, 14% and 22% of young adults (ages 19-30) reported past 30-day and past-year cannabis vaping, respectively, substantially higher than midlife adults (ages 35-50; 6% and 9%, respectively).^12^ CVPs, just like other cannabis products, may have potential applications for acute symptom relief across multiple conditions (e.g., anxiety, depression, epilepsy, pain); however, they also pose physical and mental health risks along with negative social consequences.^13–15^ Specifically, long-term and regular use of cannabis has a high potential for abuse and dependence and is associated with increased risk of paranoia, anxiety, depression, and exacerbation of psychosis.^16,17^ Among young adults, frequent cannabis use may also lead to altered brain development, poor educational outcomes, and difficulties in transition to work and adult roles.^18,19^ Although vaping cannabis reduces the ingestion of smoke-related toxins and carcinogens,^20,21^ cannabis vaping has been shown to produce a larger concentration of PM_2.5_ than tobacco cigarettes,^22^ and endothelial function impairment from exposure to CVP vapor^23^ is similar to smoked cannabis and tobacco cigarettes.^24^

One major factor contributing to CVP use may be exposure to commercial marketing. Decades of research in other areas, including alcohol and tobacco marketing, have yielded conclusive evidence that marketing exposure leads to initiation and regular use of these products.^25–29^ Youth and adults in the U.S., particularly in the states where non-medical cannabis use is legal, are exposed to cannabis advertising, including CVP-specific advertising, through many channels (e.g., physical and online retailers, billboards, websites, and social media).^30–36^ More research on CVP marketing is needed to support the development of marketing policies and other intervention strategies that can effectively prevent youth initiation and reduce use.

Marketing theories such as the means-end approach posit that advertising strategies leverage tangible *product features* detailing the functionalities and benefits of the products (e.g., a CVP that “gets me high”) to shape individuals’ expectancies (the anticipated outcomes from using the products) and interest in using the marketed products.^37,38^ These product features often appeal to consumers’ psychological needs and motives for product use. Specifically, consumers form harm perceptions and outcome expectancies of product use based on how the product features are advertised and labeled.^39–41^ Indeed, research on *nicotine* vape product marketing found that product features (e.g., claims describing flavor sensation, smoking cessation benefits, and convenience) predict and influence the positive expectancies and interest in the use of nicotine vape products among young adults.^42–47^ The CVP industry promotes CVPs as “safer” or “healthier,” especially compared with smoking cannabis products, which can reduce perceptions of harm that may delay or prevent use among young people.^48,32^ However, little else is known ssssssabout the content and types of product features used for marketing CVPs. Much of the research assessing the characteristics of cannabis product features has grouped various types of cannabis products together and only examined a narrow range of features.^49–51^ One of the few studies that limited its investigation of product marketing descriptions to a single product type (cannabis flower) revealed that the dominant product features included relaxation, stress relief, and happiness.^52^ However, product features used for promoting CVPs are likely to differ from other cannabis products due to vape products’ distinctive functions (e.g., convenience, discreetness). Evidence unique to the CVPs is greatly needed to understand how this increasingly popular cannabis product is marketed to consumers to inform policy and intervention strategies. To address this research gap, this study aimed to identify and describe the product features used for marketing CVPs from a large cannabis e-commerce marketplace.

## METHODS

### Brand and Website Selection

We first obtained a list of 100 top-selling cannabis vape brands in the U.S. in 2023 from BDSA, a global company tracking the cannabinoid sales market.^53^ To compile this list, BDSA generated the dollar sales amount of all CVP brands sold in 10 states that represent some of the largest legal non-medical cannabis markets in the country (AZ, CA, CO, FL, IL, MA, MD, MI, MO, NV, NJ, NV, NY, OH, OR, PA).^54^ The product descriptions for CVPs were obtained from Jane Technologies (www.iheartjane.com), a large e-commerce marketplace focused on the cannabis industry, featuring 22,000 cannabis brands and 1.8 million products, with over 1.6 million verified customer reviews.^55^ The website has 1.3 million visits (>95% from the U.S.) per month on average and allows consumers to order cannabis products for delivery and be connected with cannabis dispensaries near them throughout the country.^56^ The website also provides an extensive catalog of available products (including brand, product name, product descriptions, etc.) for cannabis retailers and dispensaries to create their own product inventories on their websites.^56^ The information provided in their product inventory allows customers, retailers, and the public to learn about cannabis products, including product images and customer product reviews. Product information from this online retail website is also highly accessible to young people. It is documented that close to half of the adult visitors (aged ≥18) of Jane Technologies were young adults aged 18-34.^57^ Since the visitors just need to click “confirm” for its age verification question, “Please confirm you’re over 21 or a valid medical patient” to enter the website, it is also widely accessible to underaged minors.

### Product Description Selection

Product descriptions of a sample of 343 CVPs (all regular delta-9 THC products) from 78 of the 100 top-selling cannabis vape brands – with the highest customer review ratings and recent purchases – were selected from Jane Technologies for content coding (see **Figure 1** for examples of product images and descriptions). The overall goal of the search strategy was to select CVPs with the highest customer review ratings and recent purchase history to capture the recent, positively rated products sold on this platform. First, the name of the brand from the 100 top-selling list was entered into the search bar on Jane Technologies’ home page to generate a result of all products from that brand. Then, the filters “vape” and “4+ star ratings” were applied to narrow the results down to CVPs with the highest review ratings from that brand. Finally, to further narrow down products with a recent record of product reviews, the five vape products with the most customer reviews and at least one review written since January 1, 2023, were identified. This process was repeated for all brands. In some cases, less than five products from a brand were identified because all the reviews pre-dated January 1, 2023.

**Figure 1.**
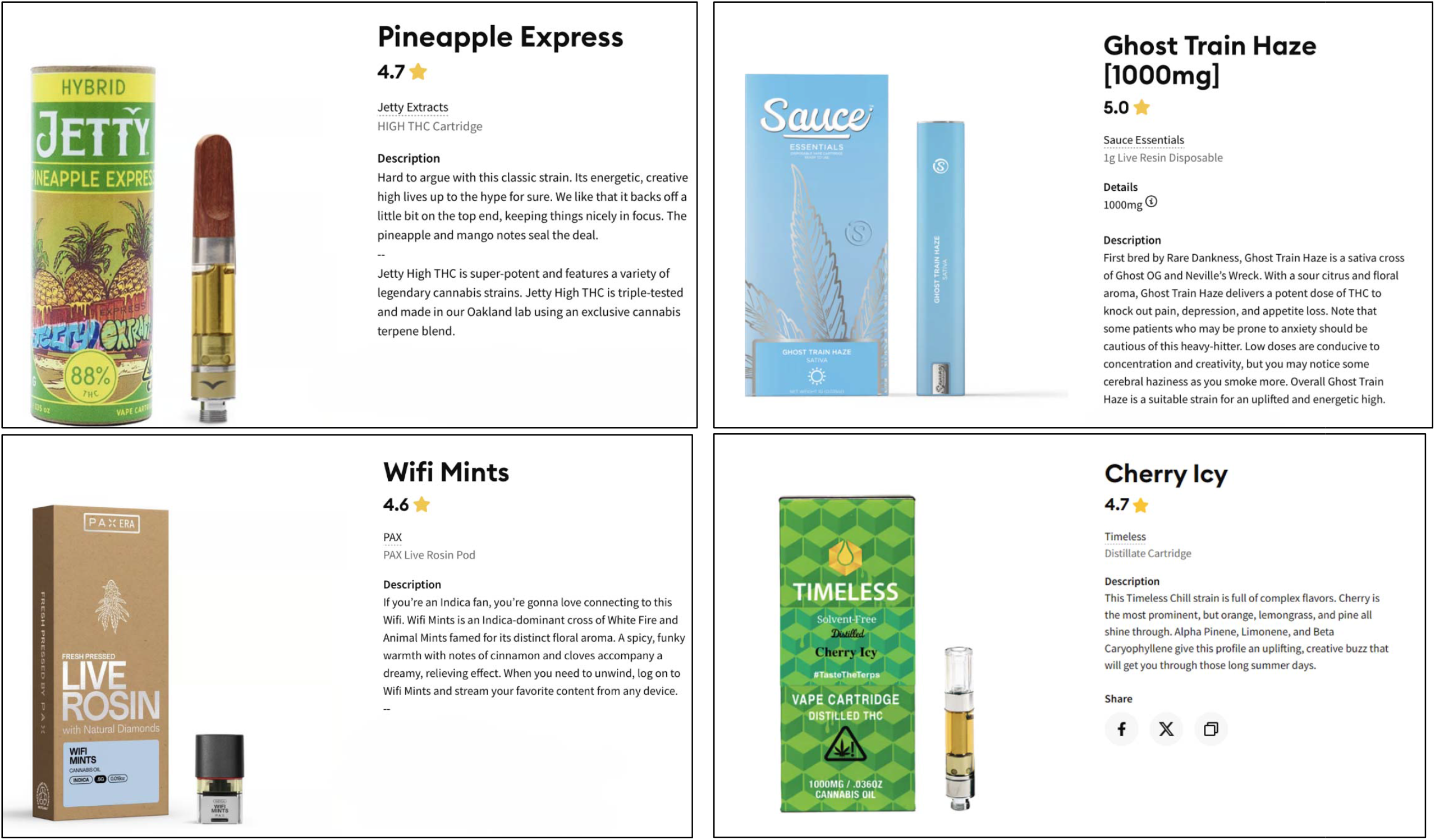
Examples of Selected Cannabis Vape Product Descriptions from a Large Cannabis E-Commerce Website.

To establish consistency in the selection of products, two members of the research team first worked on selecting CVPs from the same five brands and reconciled discrepancies in product selection. They then divided up the list of remaining brands evenly and independently selected products for those brands. We did not code the product reviews but used them to inform the selection of products only.

We did not include CVPs from 22 of the 100 top-selling brands due to at least one of the following reasons: (1) the brand did not feature any vape products on the website (*n*=10), (2) the generated products had no reviews or only had reviews written before January 1, 2023 (*n*=5), (3) the generated products had no reviews (*n*=4), or (4) the product descriptions were presented in brief bullet points that did not include adequate information related to product features for content analysis (*n*=3). After identifying the relevant CVPs that met the search criteria (*n*=343), their product descriptions were screenshotted and saved in the project folder for content coding.

### Coding Procedure

A content codebook was developed using a deductive and inductive process that relied on existing research related to individuals’ expectancies for using cannabis products in general and CVPs in particular,^51,52,58^ and a qualitative review of product descriptions to develop and generate additional codes. Codes were also created to capture nuanced product feature categories when distinct content emerged. Three coders were trained through an iterative process of reviewing the codebook and coding the descriptions. The entire research team met to discuss the created product feature codes and their broad categories. This resulted in 24 codes across 11 product feature categories: flavor profile and sensation, psychoactive effects, product quality, mood enhancement, reduced harm, relaxation/tension reduction, therapeutic effects, focus/creativity, convenience/discreetness, socialization enhancement, and physical performance enhancement. Specific codes and their definitions are listed in **Table 1**. The coders then completed two rounds of double coding using a subsample (*n*=50 each round) of product descriptions to establish inter-rater reliability and inform codebook refinement. Each product description could be coded with as many codes as applied. Discrepancies were resolved, and the codebook was refined through discussion during the team meetings. Once Krippendorff’s alpha reached >0.80 in the second coding round,^59^ the remaining product descriptions were randomly split between coders. Since many states that legalized non-medical cannabis use have marketing restrictions related to using unfounded and misleading therapeutic or health claims for marketing cannabis,^60^ we delved into these claims to assess whether they are based on the scientific evidence related to the health benefits of cannabis use^14^ (see the list of such claims in **Supplemental Table 1**). This research involved the use of publicly available data, which is not considered human subject research under 45 CFR 46.102(d) and does not require Institutional Review Board review.

**Table 1.** Product Feature Categories and Definition of Cannabis Vape Products Shown in Product Descriptions of a Large Cannabis E-Commerce Website.

| Product Feature Categories |  | Definition | Code Frequency | n | Example Product Description |
| --- | --- | --- | --- | --- | --- |
| **Flavor Profile and Sensation** |  |  | **74.1** | **254** |  |
|  | General flavor descriptor | The description includes terms used to describe the product's flavor (big flavor, flavorful, etc.) or general sensations of the product's flavor (tasty, delicious, refreshing, etc.). | 54.2 | 186 | High-quality vaporizable oils specially developed to deliver the full flavor and complexity of each strain. |
|  | Fruit flavor | The description includes the product's fruit flavor name or words that describe the product to taste like fruit(s) (citrus flavors, strawberry, watermelon, grape, etc.). | 38.5 | 132 | A balanced blend of fresh strawberries and ripe tropical bananas with undertones of sweet berries and earthy notes. |
|  | Flavor sensation profile | The description includes terms to describe the tastes, smells, and textures that make up the product's flavor, such as "sweet," "sour," and "spicy." | 30.6 | 105 | Dosi-Do is an intensely relaxing indica dominant strain, with sweet, earthy flavors and a pungent aroma that strings the nostrils. |
|  | Other flavor | The description includes the product's flavor name that is not a fruit or concept flavor or words that describe the product to taste like non-fruit related flavors such as mint, floral, dessert, candy (Wedding Cake, Mint Cookies, etc.). | 20.1 | 69 | You are immediately greeted with thee fresh from the bakery aroma of vanilla frosting and just mixed cake batter. |
|  | Concept flavor | The description includes the product's concept flavor name (Sour Diesel, Blue Dream, Jack Herer, etc.). | 11.9 | 41 | KryptoChronic is a vibrant lime green with purple undertones and orange bright pistils. Tastes and smells like you put gas in your bowl of fruity pebbles and milk. |
|  | Flavor variety | The description includes claims that the product comes in a variety of flavor options or lists the available flavor options. | 10.8 | 37 | Easy on the throat, discreet, potent, and available in a variety of flavors and strains. |
| **Psychoactive Effects** |  |  | **47.5** | **163** |  |
|  | Psychoactive descriptor | The description includes psychoactive descriptors to describe the product such as powerful, potent, or strong. | 42.0 | 144 | Jetty High THC is super-potent and features a variety of legendary cannabis strains. |
|  | Psychoactive effect | The description includes the psychoactive effects of the product on the user such as feeling high, stoned, or buzzed. | 14.6 | 50 | This hybrid delivers a well-balanced mind and body high with a juicy burst of flavor. |
| **Product Quality** |  |  | **42.3** | **145** |  |
|  | Quality descriptor | The description includes the keyword 'quality,' indicating the product is high quality. | 23.3 | 80 | The quality oil you've come to know from Select, now infused with freshly harvested live resin terpenes. |
|  | Purity descriptor | The description includes the keyword 'pure/purity/purified.' | 16.9 | 58 | Heavy Hitters Ultra Extract represents the pinnacle of cannabis purity. |
|  | Natural descriptor | The description includes the keyword 'natural.' | 14.6 | 50 | These cartridges are filled with 100% natural, solvent free CO2 extracted cannabis oil. |
|  | Premium descriptor | The description includes the keyword 'premium.' | 11.1 | 38 | Church Cannabis' product line includes super premium oil derived from the most sought-after strains. |
|  | Organic descriptor | The description includes the keyword 'organic.' | 1.5 | 5 | THC distillate, flavored with organic botanical terpenes, these vapes are mixed with flavors extracted from natural plant sources. |
| **Mood Enhancement** |  |  | **33.8** | **116** |  |
|  | Mood enhancement | The description includes claims that product use is associated with positive mood such as happiness and | 33.8 | 116 | Orange Crush is said to start with a sunny, buzzy, warmth that expands in waves throughout your body, |
|  |  | euphoria, or associated with avoiding negative mood such as feeling down, coping, etc. |  |  | leaving you energized, care-free, and vibrating with good vibes. |
| **Reduced Harm** |  |  | **24.2** | **83** |  |
|  | Reduced harm | The description includes terms that indicate products have low or reduced harm, including metal-free, safe, safety, no detection of chemicals or add-ons. | 24.2 | 83 | An uncompromising oil that delivers – without additives, artificial terpenes, or cutting agents. |
| **Relaxation/Tension Reduction** |  |  | **23.9** | **82** |  |
|  | Relaxation/tension reduction | The description includes terms or phrases to describe the relaxing effects of product use such as relaxed, calm, or chill, etc. | 23.9 | 82 | The strong relaxation can be felt throughout the entire body and is often reported to be used for soothing, tranquil relief. |
| **Therapeutic Effects** |  |  | **18.4** | **63** |  |
|  | Mental health | The description includes health-related information regarding claims of therapeutic benefits for specific <u>mental health issues</u> such as stress, anxiety, depression, etc. | 7.6 | 26 | Smokers often use this strain for alleviation of anxiety and stress. |
|  | Health lite | The description includes non-clinical and non-specific but health-related claims and words, such as therapeutic, promoting wellness, treatment, remedy, etc. | 6.7 | 23 | Matter offers the highest level of safe, effective, and consistent therapeutic relief. |
|  | Physical health | The description includes health-related information regarding claims of therapeutic benefits for specific <u>physical health issues</u> such as pain, cancer, arthritis, multiple sclerosis, etc. | 5.3 | 18 | The cerebral effects of this strain eventually lend to a palpable sense of physical relaxation that may aid in pain management throughout the day. |
|  | Sleep | The description includes claims of sleep benefits from product use, such as improving sleep or helping with insomnia. | 5.0 | 17 | The perfect strain to provide a euphorically-sleepy, heavy high. |
| **Focus/Creativity** |  |  | **16.6** | **51** |  |
|  | Focus/creativity | The description includes claims that product use will result in better focus or concentration, alertness, creativity, awareness, etc. | 16.6 | 51 | The Sativa dominance of this cartridge can provide a clear-headed and focused experience, making it a great choice for activities that require mental clarity, such as brainstorming or creative work. |
| **Convenience/Discreetness** |  |  | **11.1** | **38** |  |
|  | Convenience/discreetness | The description includes claims that the product is convenient or discreet to use, such as being easy to conceal and use on the go. | 11.1 | 38 | It not only offers a discreet vaping experience, but the flavor will keep you coming back for more! |
| **Socialization Enhancement** |  |  | **6.7** | **23** |  |
|  | Social effects | The description includes claims that product use may help with friendship, familiarity, romance, and closeness with others. May also include suggestions of using the product at social events (happy hour, parties, etc.). | 6.7 | 23 | More discreet than a joint, and faster acting than edibles, they're the perfect accessory for moments with friends. |
| **Physical Performance** |  |  | **6.1** | **21** |  |
|  | Physical performance | The description includes claims that product use is associated with physical improvement effects such as physical strength, energy, and ability (e.g., exercising, singing, sexual performance). | 6.1 | 21 | This creative, energetic strain could be very useful for a person to sing, dance, and chat with friends at ease. |

## RESULTS

**Table 1** presents the frequencies of product features found in the sample of CVP descriptions as well as detailed descriptions, definitions, and examples of those product features. The most frequent product feature category was flavor profile and sensation, referenced in 74.1% of descriptions. This category included general flavor descriptors (54.2%), fruit flavors (38.5%), flavor sensation profile (30.6%), other flavors (20.1%), concept flavors (11.9%), and flavor variety (10.8%). Psychoactive effects appeared in about half of the descriptions (47.5%) and included references to product potency (42.0%) and psychoactive effects on the user (14.6%). Product quality appeared in 42.3% of the descriptions, comprising content related to quality (23.3%), purity (16.9%), natural (14.6%), premium (11.1%), and organic (1.5%). Other product features promoted were mood enhancement (33.8%), reduced harm (24.2%), relaxation/tension reduction (23.9%), and therapeutic effects (18.4%). Therapeutic effects included mental health benefits (7.6%), non-clinical health-related content referred to as health lite (6.7%), physical health benefits (5.3%), and sleep benefits (5.0%). Less commonly promoted product features were focus/creativity (16.6%), convenience/discreetness (11.1%), socialization enhancement (6.7%), and physical performance enhancement (6.1%).

## DISCUSSION

This study offers insights into how the cannabis industry promotes CVPs’ functions and benefits, which is crucial given the influence of these features on consumers’ perceptions and interest in using the products.^37,38^ This study identified 11 product feature categories used to promote top-selling and highly rated CVPs on a large cannabis e-commerce marketplace. The results can promote policy advances and corrective messaging to counteract the influence of the cannabis industry’s marketing strategies for promoting CVPs, a type of cannabis product increasingly gaining market share and popularity, especially among young people.

The most common product feature category promoted was flavor profile and sensation, which has previously been shown to be among the most frequently used product features in other cannabis marketing content analyses.^52,61,62^ Particularly salient flavor descriptors included “flavorful,” “delicious,” and “tasty,” which suggest sensory appeal. Fruit flavors were the most marketed CVP flavors in our study. Flavors in general, and fruit flavors in particular, may be especially enticing to younger people, as suggested by mounting literature related to the marketing influence of flavored tobacco products, especially flavored nicotine vape products.^43,45,46^ Further, research has shown that many consumers who vape cannabis, especially younger people, use flavored CVPs that taste like fruit, candy, or dessert,^63^ and flavors serve as an important reason for CVP initiation and susceptibility.^64^ Therefore, more research is needed to investigate how product features highlighting flavor sensations and specific youth-appealing flavors may promote young people’s product use to further inform marketing and labeling policies on cannabis flavors.

Psychoactive effects are another frequently used product feature for promoting CVPs. Other content analyses^51,52^ found that including claims and messages about the psychoactive effects of cannabis products is a common promotional tactic on cannabis retailer websites. Viewing psychoactive effect claims may heighten the intention to use the promoted products, especially because “getting high” is found to be a common motivation and reason for cannabis use.^65,66^ Additionally, psychoactive effect features may be particularly influential for those who are interested in using CVPs as consumers reported perceiving CVPs to produce stronger, quicker, and more favorable “highs” compared to other modes of cannabis.^67,68^ Recent studies have also found that the concentrates used in CVPs are much more potent than other dominant cannabis products, such as smoked products,^69,70^ potentially increasing the users’ risks of developing cannabis dependence.

We also found a large collection of quality-related product descriptors including the terms “quality,” “purity,” “natural,” “premium,” and “organic.” Consumers might interpret products with those claims as healthy or less harmful. Product quality descriptors are frequently used in tobacco marketing^71^ and can lower consumers’ perception of the tobacco products’ harm and escalate interest in using the products.^72,73^ This phenomenon is similar to the “health halo” effect observed in food and tobacco industry marketing^74,75^ where consumers may overestimate a product’s healthiness based on ingredient claims suggesting the quality or cleanness of the products. We also found several product feature claims that may directly convey reduced risks and harm from using CVPs, including “chemical free,” “no additives,” or “non-GMO.” This finding is consistent with a previous content analysis^61^ that found healthy ingredient descriptors (e.g., “non-GMO,” “artificial free”) to be a common type of product feature on the front of cannabis edible packages. It is concerning that these two categories of product features combined appear in more than 70% of product descriptions from our content analysis, indicating these marketing materials’ potential impact on reducing individuals’ perceived harm from CVP use. Research has already shown that many consumers view CVPs as less harmful compared to other methods of cannabis use, such as smoking.^68,76^ With the potential legalization of cannabis in more states, consumers may increasingly view cannabis or CVPs as not harmful or risky.^77,78^ More research is needed to examine the impact of viewing those claims on perceived risk and harm from CVP use to inform marketing and labeling restrictions and corrective messaging.

In addition, although cannabis was initially legalized by many states in the U.S. for its medical effects, less than 20% of the products featured claims relevant to the therapeutic effects of CVP use. These therapeutic effect claims were comprised of content about health in general (e.g., “promotes wellness”), mental health (e.g., “reduces anxiety and depression”), physical health (e.g., “aids in pain management”), and sleep benefits (e.g., “helps with insomnia”). A deeper delve into the content of those claims, however, revealed that some of them may be unfounded and misleading to consumers, especially related to the treatment of mental health conditions, which can lead to product misuse.^14,79^ For example, we found more than 40% of therapeutic claims were relevant to mental health conditions (e.g., anxiety and depression) and their treatment. However, substantial evidence has shown that frequent use of cannabis may increase the chance of developing mental health conditions such as anxiety^80^ and psychosis.^81,82^ This use of unfounded claims may further increase the risk that those who have mental health conditions such as depression and anxiety will use the products for symptom relief, which may exacerbate their condition. Several therapeutic claims targeted specific populations, such as patients and smokers, which may convey unfounded health benefits of CVP use among these groups who already have increased health risks. The U.S. Federal Trade Commission’s (FTC) truth-in-advertising law prohibits deceptive advertising claims regarding the safety, efficacy, and other benefits of health-related products, including cannabis.^83^ According to the FTC’s Health Product Compliance Guideline, companies must support health-related claims with high quality evidence.^84^ The FTC has taken action against companies that have failed to meet these guidelines, including sellers of cannabidiol (CBD) products promoted with unsupported health claims.^85^ Such monitoring of therapeutic claims used in CVP marketing materials is essential to prevent misleading information that may impact consumers’ decisions. Additional research is greatly needed to assess how these claims are interpreted by and influence the public, including those with increased health risks, to inform product marketing and labeling policies.

Many other functions and benefits found in our content analysis, such as mood enhancement, socialization enhancement, and focus/creativity, are specifically matched to cannabis use expectancies well established in previous literature.^86–89^ For example, mood enhancement included claims that using the product would provide positive feelings, such as euphoria or happiness; and socialization effects included claims that product use may help with friendships, romance, and closeness to others. Therefore, according to the advertising theory of the means-end approach, which posits that product features influence product use by first shaping individuals’ motives and outcome expectancies,^37,38^ these product features might be especially effective in promoting use interest. Research is greatly needed to investigate how= those product feature claims may affect specific cannabis-related expectancies, which may further drive product use intentions and behaviors.

Despite the vast categories of product features in place to promote CVPs, we did not find any warnings or relevant information about health risks from using CVPs that may serve to prevent product use or misuse. It has been shown that viewing either visual or textual warnings about cannabis products is associated with an increase in knowledge about health effects^90^ and a decrease in product appeal^90,91^ and use intentions.^91^ Providing such information can help potential consumers, especially underaged minors and those who already have physical and mental health conditions, make informed decisions when considering purchasing and using the products. Regulations that mandate warnings about potential health risks and side effects on cannabis marketing channels, especially cannabis retailer channels where consumers can directly purchase the products and with soft age gating, are greatly needed to help reduce product misuse.

The current analysis has several limitations. First, we analyzed the content of product descriptions from top-selling CVP brands with the most favorable customer reviews and recent purchase history based on the information from only one cannabis e-commerce website. Therefore, we cannot be sure that the sample of product descriptions is fully representative of CVPs of the same characteristics available in the current marketplace. Second, cannabis e-commerce websites are more likely to be visited by existing consumers of cannabis who already have purchase intentions or have used the products before. Young people who do not use cannabis, a priority population for non-medical cannabis use prevention, may be exposed to CVP marketing materials from other marketing channels (e.g., outdoor billboards and social media) that may include different product features. Lastly, this content analysis did not include information related to the packaging of CVPs, which may also contain content features that suggest product functions and benefits, such as images and text related to the flavors and therapeutic and psychoactive effects.

## CONCLUSIONS

This content analysis revealed a series of product features used for marketing popular CVPs on a leading cannabis retail website. Some of these product features, such as flavor profile and sensation and psychoactive effects, may be especially appealing to youth. Some other frequently used product features, such as product quality and reduced harm, may mislead consumers to perceive the products as low-risk or healthy to use. Many product features identified are distinctly matched to various cannabis use outcome expectancies, potentially exerting a strong influence on individuals’ interest in using the products. Additional research is needed to understand the impact of promoting these product features in commercial CVP advertising to inform policy advances on product marketing and labeling, as well as corrective messaging to reduce the impact of viewing those product features on initiating and increasing product use.

## Data Availability

All data produced in the present study are available upon reasonable request to the authors

---

**Supplemental Table 1.** Therapeutic Effect Claims for Promoting Cannabis Vape Product Descriptions a Large Cannabis E-Commerce Website.

| **Therapeutic Effect Claims** | **Health Lite** | **Mental Health** | **Physical Health** | **Sleep** |
| --- | --- | --- | --- | --- |
| Patients report swift balancing and refreshing effects, making it popular to enjoy during the day for calm, focus, and uplifting relief. | X |  |  |  |
| Patients report enjoying GDP for calming relief while conveying gentle mental stimulation, excellent for relaxation without overt sedation. | X |  |  |  |
| Patients report invigorating and uplifting cerebral effects, making it great for relief when you need to be active. | X |  |  |  |
| Smokers often use this strain for alleviation of anxiety and stress. |  | X |  |  |
| Lemon Pound Cakes' initial effects are more cerebral, bringing an uplifting mental state and clarity to the mind after even the most stressful of days! |  | X |  |  |
| OG's terpene profile provides some of the most relaxing and medicinal effects for those looking for sleep and pain relief. | X |  | X | X |
| Ideal for a true indica connoisseur, this dense bud packs a strong body high that melts away stress, pain, and anxiety. Whether you're managing pain, nausea, insomnia, or just trying to relax, Ice Cream Cake is an effective, appetizing strain that will never fail to make you come back for seconds. |  | X | X | X |
| Large buds ripple with bright orange hairs containing a citrusy, herbal flavor that lifts up the mind while relieving it of stress and anxiety. |  | X |  |  |
| Indulge in a creamy, sweet, and soothing hybrid, and let Wedding Cake carry you off to bed. |  |  |  | X |
| Guava 2.0's effects are indica-forward, which makes for an effective strain when combating stress and pains. |  | X | X |  |
| The cerebral effects of this strain eventually lend to a palpable sense of physical relaxation that may aid in pain management throughout the day. |  |  | X |  |
| Ideal for pain relief, this strain has a soft onset with strong physical relaxing effects. |  |  | X |  |
| Its effect is mildly energetic yet clear headed, perfect for patients looking to enjoy a connoisseur-grade strain while avoiding fatigue. | X |  |  |  |
| Sleep tonight. Friends tomorrow. |  |  |  | X |
| We pride ourselves at Grow Sciences on our ability to produce quality solventless extracts for Arizona dispensaries and our patients. | X |  |  |  |
| The perfect strain to provide a euphorically sleepy, heavy high. |  |  |  | X |
| Formulated for health and wellness, these CBD-rich blends deliver consistent effects and flavor from one batch to the next, at an affordable price. | X |  |  |  |
| With myrcene, limonene, and caryophyllene, patients report less anxiety and a noticeable breakthrough in the fog, rounding out a tasty and pleasant experience. | X | X |  |  |
| As an indica-dominant hybrid, Ghost OG can melt all your worries, stressors, and anxieties away and replace them with a soothing, peaceful experience. This can help combat feelings of pain, anxiety, nausea, and inflammation. |  | X | X |  |
| This energizing blend is full of surprises and could help relieve anxiety and pain. Illicit Sciences distillate is for those that crave a little flavor and fun with their method of medicating. | X | X | X |  |
| Those who consume Chrome Slipper 99 may experience feeling creative, happy, hungry, relaxed, and sleepy. |  |  |  | X |
| Cannalope Kush is prized by cannabis enthusiasts for its balanced effects, delightful flavor profile, and potential therapeutic benefits. | X |  |  |  |
| As you inhale the earthy and pungent flavors, you can feel the stress and tension of the day melting away, replaced |  | X |  |  |
| by a soothing and relaxing sensation. |  |  |  |  |
| As you inhale the sweet and cheesy flavors, you can feel the stress of the day melting away. |  | X |  |  |
| Matter offers the highest level of safe, effective, and consistent therapeutic relief to people suffering from chronic medical conditions. |  | X | X |  |
| Its effects may help with managing nausea, stress, minor body aches, and sleeplessness. |  | X | X | X |
| Find your ‘feel’ (energizing, euphoric, sleepy) and pick a flavor. |  |  |  | X |
| Providing a flavorful escape from stress, anxiety, and depression. |  | X |  |  |
| Its strong physical effects and uplifting mental high make it a perfect end-of-the-day strain and a nice match for folks contending with stress, restlessness, and pain. |  | X | X |  |
| Forbidden Fruit’s deep physical relaxation and mental stoniness make it perfect for dulling minor physical discomfort and discarding stress. |  | X | X |  |
| Our Indica is perfect for those days spent curled up on a couch, a soothing relaxing evening or getting a restful night’s sleep. |  |  |  | X |
| The Blissful Berry terpene profile promotes an indica dominant sensation, offering a relaxing body euphoria great for melting away muscle tension, stress, and anxiety. |  | X | X |  |
| This terpene profile is best for those looking for relief from inflammation, pain, stress, and anxiety. |  | X | X |  |
| Users can expect relief from insomnia and reduced anxiety. |  | X |  | X |
| Users can expect pain relief, creativity, and a sense of euphoria. |  |  | X |  |
| The high THC content will awaken your senses and may relieve symptoms of pain, depression, and nausea. |  | X | X |  |
| It provides great taste and carefree relaxation that can help soothe pain, stress, and anxiety. |  | X | X |  |
| ROVE is pumped to offer this feisty sativa – good for managing stress, minor body aches, and nausea. |  | X | X |  |
| Ghost Train Haze delivers a potent dose of THC to knock out pain, depression, and appetite loss. |  | X | X |  |
| Strawberry Cough is a great solution in times of elevated stress. |  | X |  |  |
| The combination results in a high extraction oil, available in plenty of strain options to meet a wide range of patient needs. | X |  |  |  |
| Available in a variety of strains to suit a wide range of patient needs. | X |  |  |  |
| The effects of this daytime favorite tend to be tasty as well, beginning with a boost of energy and focus, followed by all-over relaxation and light sedation. |  |  |  | X |
| Strain-specific oil or ultra-refined distillate oil are used to deliver all the therapeutic benefits of our best-selling Verano flower via food-grade ethanol extraction in convenient disposable pen. | X |  |  |  |
| While many choose Strawberry Cough for its flavor, they come back for its effects: an uplifting euphoria that gently fades feelings of stress and depression. |  | X |  |  |
| Kewl-aid for grown-ups, Wavelength’s Purple Punch cartridges make taking medicine fun. | X |  |  |  |
| Drift off to sleep under a sky of wavy lights with this budtender favorite. |  |  |  | X |
| The blend of alpha-humulene and beta-caryophyllene relaxes the body, easing away stress. |  | X |  |  |
| Stress less and enjoy more, the oil in this 510-cart pairs nicely with a glass of wine and a beautiful sunset. |  | X |  |  |

